## Supplementary Information for "An imperfect tool: contact tracing could provide valuable reductions in COVID-19 transmission if good adherence can be achieved and maintained"

### Transmission parameter distributions

The generation interval is the time difference between a case being infected and that case infecting others, whereas the serial interval is the time difference between the onset of symptoms in the primary case and the secondary case. As such, the generation interval is, by definition positive, whereas the serial interval can be negative in cases where transmission occurred during the primary case's pre-symptomatic period. Figure S1 shows the fitted distributions for the incubation period, transmission density profile, serial interval and generation interval used in the model.

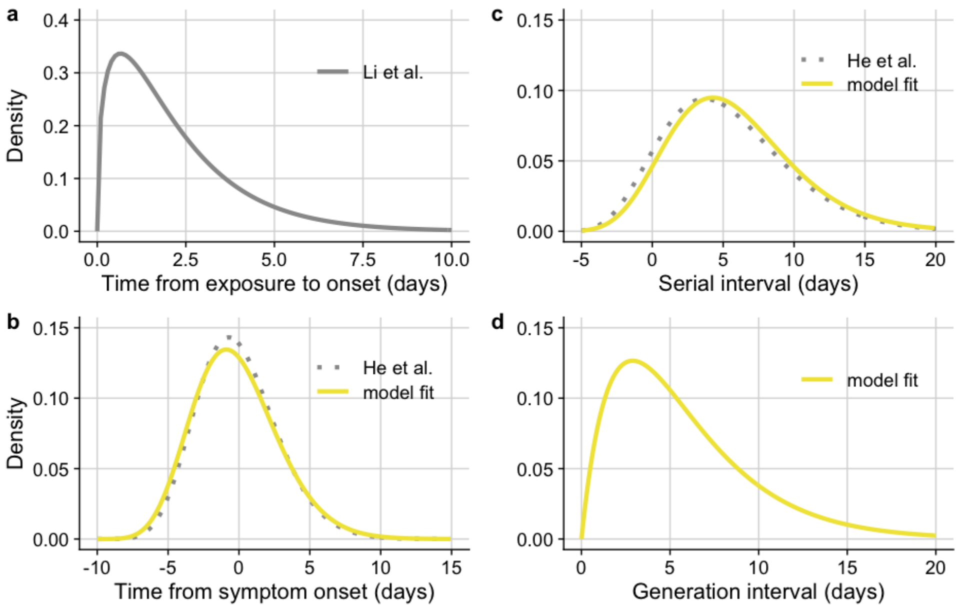


Figure S1: **Transmission parameter distributions.** Distributions for a) incubation period (exposure time to symptom onset) from Li et al. [1]; b) transmission profile relative to symptom onset, fitted to data and compared to He et al. [2]; c) serial interval, fitted and compared to He et al. [2]; and d) generation interval, combined distribution from a) and b) with re-sampling to prevent negative serial intervals, as described in the main text.

### CMMID working group authorship list

The following authors were part of the Centre for Mathematical Modelling of Infectious Disease 2019-nCoV working group. Each contributed in processing, cleaning and interpretation of data, interpreted findings, contributed to the manuscript, and approved the work for publication: Alicia Rosello, Rosalind M. Eggo, Quentin J Leclerc, Matthew Quaife, Emily S. Nightingale, Thibaut Jombart, Jon C. Emery, Timothy W. Russell, Nikos I. Bosse, Megan Auzenbergs, Amy Gimma, Charlie Diamond, Yang Liu, Kiesha Prem, Eleanor M. Rees, Christopher I. Jarvis, W. John Edmunds, Akira Endo, David Simons, Billy J. Quilty, Damien C. Tully, Oliver Brady, C. Julian Villabona-Arenas, Gwenan M. Knight, Samuel Clifford, Stéphane Hué, Kevin van Zandvoort, Adam J. Kucharski, Mark Jit, Arminder K. Deol, Kathleen O'Reilly, Anna M. Foss, Fiona Yueqian Sun, James D. Munday, Hamish P. Gibbs, Rein M. G. J Houben, Rachel Lowe, Sophie R. Meakin, Georgia R. Gore-Langton, Sebastian Funk, Carl A. B. Pearson, Katherine E. Atkins, Simon R. Procter, Stefan Flasche, Nicholas G. Davies.

The following funding sources are acknowledged as providing funding for the working group authors. Alan Turing Institute (AE). BBSRC LIDP (BB/M009513/1: DS). This research was partly funded by the Bill & Melinda Gates Foundation (INV-003174: KP, MJ, YL; NTD Modelling Consortium OPP1184344: CABP; OPP1180644: SRP; OPP1183986: ESN; OPP1191821: KO'R, MA). DFID/Wellcome Trust (Epidemic Preparedness Coronavirus research programme 221303/Z/20/Z: CABP, KvZ). Elrha R2HC/UK DFID/Wellcome Trust. This research was partly funded by the National Institute for Health Research (NIHR) using aid from the UK Government to support global health research. The views expressed in this publication are those of the authors and not necessarily those of the NIHR or the UK Department of Health and Social Care (KvZ). ERC Starting Grant (757688: CJVA, KEA; 757699: JCE, MQ, RMGJH). This project has received funding from the European Union's Horizon 2020 research and innovation programme - project EpiPose (101003688: KP, MJ, WJE, YL). This research was partly funded by the Global Challenges Research Fund (GCRF) project 'RECAP' managed through RCUK and ESRC (ES/P010873/1: AG, CIJ, TJ). HDR UK (MR/S003975/1: RME). Nakajima Foundation (AE). NIHR (16/137/109: BJQ, CD, FYS, MJ, YL; Health Protection Research Unit for Modelling Methodology HPRU-2012-10096: NGD, TJ; PR-OD-1017-20002: AR). Royal Society (Dorothy Hodgkin Fellowship: RL). UK DHSC/UK Aid/NIHR (ITCRZ

03010: HPG). UK MRC (LID DTP MR/N013638/1: EMR, GRGL, QJL; MC_PC 19065: RME; MR/P014658/1: GMK). Authors of this research receive funding from UK Public Health Rapid Support Team funded by the United Kingdom Department of Health and Social Care (TJ). Wellcome Trust (206250/Z/17/Z: AJK, TWR; 206471/Z/17/Z: OJB; 208812/Z/17/Z: SC, SFlasche; 210758/Z/18/Z: JDM, NIB, SFunk, SRM). No funding (AKD, AMF, DCT, SH).
